# Nurses’ Knowledge, Attitudes, and Practices on Sedation Practices for Patients Admitted to the Intensive Care Unit at Tertiary Hospitals in Dar es Salaam

**DOI:** 10.64898/2026.01.21.26344577

**Authors:** Ibrahim M. Luge, Masunga K. Iseselo, Sofia Sanga

## Abstract

**Background:** Sedation is a medically induced state implemented to facilitate procedures or care of critically ill patients in the Intensive Care Unit (ICU). Nurses ensure safe sedation by applying agitation scales to reduce complications. There is sufficient research evaluating nurses’ knowledge, attitudes, and practices regarding sedation for critically ill patients in the ICU of Tanzanian tertiary Hospitals.

**Methods:** This hospital-based cross-sectional study assessed nurses’ knowledge, attitudes, and practices on sedation in ICUs of three referral hospitals in Dar es Salaam, Tanzania. A sample of 163 nurses was recruited using stratified sampling. Data were collected through structured questionnaires and checklists, then analyzed with SPSS version 25. Variables with p ≤ 0.2 entered multivariate regression; significance was set at p < 0.05, 95% confidence interval.

**Results:** Most participants were female (60.7%) with a mean age of 34.6 ± 6.48 years. Of 163 nurses, 68.1% had adequate knowledge of sedation, though males were 38% less likely to demonstrate this (AOR = 0.377, 95% CI: 0.181–0.787, p = 0.009). Unfavourable attitudes were reported by 76.7% (n=125), while trained nurses were nearly twice as likely to show favorable attitudes (AOR = 2.53; 95% CI: 1.015–6.422; p = 0.046). Poor adherence was noted in 81.6% of respondents. Nurses aged 25–35 were 34% less likely to report poor adherence than those aged 45 and above (AOR = 0.344; 95% CI: 0.129–0.912; p = 0.032).

**Conclusion:** This study identified adequate knowledge but negative attitudes and poor adherence among ICU nurses. Strengthening training, mentorship, and standardized protocols is crucial for enhancing sedation care and improving patient outcomes.

## Introduction

The Intensive Care Unit (ICU) provides specialized, life-sustaining care for patients with life-threatening conditions (1). Sedation is an indispensable intervention, frequently used to help patients tolerate invasive procedures, particularly mechanical ventilation; evidence suggests that up to 92% of patients undergoing mechanical ventilation receive sedatives (2). Effective sedation offers crucial benefits by managing patient distress, reducing anxiety, and alleviating pain perception (3). Sedation management is, therefore, a central component of ICU care for critically ill patients. ICU Nurses play a pivotal and independent role in administering and monitoring sedatives, directly influencing patient outcomes and recovery (4). The need for standardized care led to the publication of the Clinical Practice Guidelines for Managing Pain, Agitation, and Delirium (PAD) in adult ICU patients, which establishes the framework for safe practice (5).

Safe sedation requires the consistent use of validated sedation-agitation scales both before and after sedative administration (3). These scales must be expert-approved, reliable, and used strictly in accordance with established guidelines (5). Unfortunately, global data indicate that the utilization of these essential sedation-agitation scales remains significantly limited, despite numerous expert recommendations (6). This failure to consistently adhere to guidelines manifests in clinically risky outcomes, such as over-sedation risk. A Malaysian study indicated that 58% of ICU patients were deeply sedated during the first 48 hours and beyond (7). Such over-sedation, which can only be identified through proper scale use, is linked to prolonged ventilation and recovery times. Adverse Outcomes in the USA, a survey of Pediatric ICU (PICU) nurses reported that 31.8% of children were over-sedated, and 29.4% of critically ill PICU patients had an increased length of stay directly attributable to inappropriate sedation practices (8). These findings underscore a persistent gap in core sedation nursing practices.

Inconsistencies in the knowledge, attitude, and practice (KAP) of ICU nurses regarding safe sedation are evident worldwide, but particularly pronounced in the African context: East Africa: A 2022 study in Kenya highlighted a low rate of adherence, finding that only 37 out of 89 ICU nurses consistently used sedation-agitation scales to monitor patients (9). This signals a clear deficiency in the routine application of critical monitoring tools. North Africa: A 2023 survey of Egyptian ICU nurses exposed critical inadequacies in foundational competencies: 94.3% of nurses had an unsatisfactory basic knowledge of sedation, and 89.5% lacked adequate knowledge of sedation assessment and management (10). Furthermore, 73.3% of these nurses demonstrated incompetent practices in administering sedative medications, underscoring a profound need for targeted education and training (10).

Therefore, this study aimed to assess the knowledge, attitudes, and practices of ICU nurses regarding sedation for critically ill patients in tertiary hospitals in Dar es Salaam. The gap was poor adherence to evidence-based sedation practices among ICU nurses, and it is important because it directly affects patient safety, recovery, and quality of critical care.

## Materials and methods

### Study design

This was a cross-sectional hospital-based study aimed to assess knowledge, attitudes, and practices among nurses working in ICUs at tertiary hospitals in Dar es Salaam. Muhimbili University of Health and Allied Sciences Research Ethical Committee approved this study. A quantitative research approach was most suitable for this study because it enabled the measurement of specific variables, statistical analysis, and the identification of patterns and relationships within the data (11).

### Study setting

The study was conducted at Muhimbili National Hospital sites (MNH Upanga and Mloganzila), the Jakaya Kikwete Cardiac Institute (JKCI), and the Muhimbili Orthopaedics Institute (MOI), which are National Referral Hospitals in Dar es Salaam. The focus was on the Intensive Care Unit (ICU) and Highly Dependent Unit (HDU), where patients receive specialized treatment.

### Study population

The study population comprised all registered nurses currently employed across the identified Intensive Care Units (ICUs) and High Dependency Units (HDUs) of the participating hospitals. The total target population consisted of 240 registered nurses who were considered eligible based on a minimum criterion of six months of clinical experience in their respective units. This inclusion threshold ensured that all participants possessed sufficient exposure to routine sedation practices, making them relevant subjects for assessing current knowledge, attitudes, and practices (KAP). The final analysis focused on nurses who were available and consented to participate during the designated data collection period.

### Sampling method

A stratified proportional random sampling technique was employed to ensure that the final sample was representative of the broader nursing population across the selected institutions. Stratification was based on hospital and unit type (ICU & HDU), allowing proportional allocation of participants from each stratum. This approach enhanced the generalizability of the findings, minimised selection bias, and ensured that diverse experiences and perspectives were captured without surveying the entire population.

### Sample size estimation

The required sample size was calculated using the formula for a finite population, given the known population size (N = 163). The calculation was based on a 95% confidence level (Z = 1.96), a 5% margin of error, and an anticipated population proportion (p) of 50%, as derived from a previous study in Ethiopia (12). This yielded a minimum sample size of 148. To ensure adequate statistical power, a 10% non-response rate was added, resulting in a final target sample size of 163 nurses.

### Data collection tools and procedures

The study included adult registered nurses working in Intensive Care Units (ICUs) and High Dependency Units (HDUs) at tertiary national hospitals in Dar es Salaam. All eligible nurses received detailed verbal and written information about the study objectives, procedures, potential risks, and benefits. Participation was entirely voluntary, and written informed consent was obtained from each participant prior to enrollment, as the study involved direct participation through questionnaire completion and structured observation of clinical practice. Written consent was required to ensure that participants explicitly agreed to take part and understood their rights, including the right to decline or withdraw without any professional consequences. Data collection was conducted between 01^st^ May 2025 and 05^th^ July 2026.

Data were collected using structured self-administered questionnaires and observational checklists. The questionnaires were distributed in paper format and included clear instructions to guide participants during completion. This approach allowed nurses to complete the questionnaires at a convenient time during their shifts, minimizing disruption to routine clinical duties.

Sedation practices were assessed through structured direct observation using a predefined checklist. Trained observers systematically recorded nurses’ sedation-related practices during routine clinical care. Observations were conducted across different shifts (day and night) to capture variations in practice and ensure comprehensive assessment. Upon completion, all questionnaires and observation checklists were collected, coded, and securely stored by the principal investigator to maintain data integrity and confidentiality.

### Variables and outcome measures

Assessed nurses’ knowledge, attitude, and practice on sedation via an 8-item questionnaire covering protocols, pharmacology, and RASS/SAS scores. Defined ≥75% as adequate knowledge; ≤74% as inadequate (13). Evaluated nurses’ attitudes toward ICU/HDU sedation using a six-item Likert scale (1 = Strongly Disagree to 5 = Strongly Agree). Responses converted to percentages and classified as favorable (>50%) or unfavorable (<50%) (14) & (15). Practices were evaluated via an 8-item checklist, with adherence classified according to the cut point for good adherence (>80%) or poor adherence (<80%). Used an eight-item checklist and observation sheet during nurses’ shifts to evaluate sedation practice, scoring each item as YES or NO (16) & (17).

Categorized independent variables as education level (Diploma, Bachelor’s, Master’s) and experience duration (<5 years, >5 years) (6). The World Health Organization (WHO) supports using workload-based staffing tools like WISN (Workload Indicators of Staffing Need) to determine appropriate staffing norms, especially in resource-limited settings. These tools help tailor staffing to actual patient needs rather than fixed ratios alone (18). Nurse-patient ratios are calculated as continuous variables. The number of patients per nurse is represented by the nurse-patient ratio during a typical shift, treated as a continuous variable (e.g., 1 nurse for 1 patient =1, 1 nurse for 2 patients = 0.5, 1 nurse for 4 patients = 0.25) (6). Analyzed benchmarked relationships between independent variables and nurses’ knowledge, attitudes, and practices in ICU sedation management.

Adapted and refined prior questionnaires and checklists to align with the study’s objectives, criteria, and contextual requirements (19). Overall, structured questionnaire questions used to assess knowledge and attitudes ensured a robust and rigorous approach to participant selection and data collection. Sedation practice was assessed via a checklist based on day and night shift observation (20). Data collected over three weeks; distributed questionnaires followed up to ensure return of completed forms. The study employed a mixed-methods design to assess ICU nurses’ sedation practices. Behavioral data were acquired via scheduled, direct observations using a structured checklist and workflow sheets, ensuring temporal consistency and capturing contextual factors(21). Concurrently, cognitive data (knowledge and attitudes) were gathered via a self-administered questionnaire for systematic evaluation (22), . This multi-modal approach ensured high scientific rigor and ecological validity. All instruments were meticulously collected and stored to maintain data integrity and confidentiality.

### Trustworthiness

The data collection instruments’ validity was ensured through pre-testing with critical care specialists and subsequent revisions. Internal consistency reliability was confirmed using Cronbach’s Alpha (α) on the pre-test data, with all domains exceeding the threshold of 0.70. This rigorous process guarantees the quality of the data, providing a solid foundation for assessing nurse KAP.

### Data management and analysis

The cleaned data were subsequently coded and entered into a computer using the Statistical Package for Social Sciences (SPSS), version 25. All independent variables from the bivariate analysis with a p-value less than 0.25 were included in the model (23). Variables with an adjusted odds ratio and a p-value < 0.05 at a 95% confidence interval were deemed statistically significant in their association with knowledge, attitudes, and practices related to sedation (23)

### Ethical Considerations

Ethical approval was obtained from the MUHAS Senate of Research and Ethical Committees with reference number DA.282/298/01. C/2822 before conducting the study, after submission of the proposal. The permission to conduct this study was attained from the hospital research and administration committee at MNH Upanga, MOI, JKCI, and MNH Mloganzila. Written informed consent was obtained from each study participant voluntarily, and they were free to withdraw from the study at any time. Each study participant was informed about the study’s aims, the procedures for completing the questionnaire, the benefits of participating, and any potential risks.

Participants were identified by code number, which appeared at the top of the questionnaire paper to ensure confidentiality. Additionally, data collection took place in a private room, where a researcher and one study participant were present to maintain privacy. The study participants were told that the risk of this study was the time consumed while filling out a questionnaire. The complete filled questionnaires were collected, arranged, and stored well in the file to which only the principal investigator was the one who had access to ensure confidentiality.

## Results

The study achieved a 100% response rate, with all 163 participants successfully enrolled. The majority were aged between 25 and 35 years, with a mean age of 34.6 years and a standard deviation (SD) of ± 6.48 years; the majority of the study respondents, 60.7% (99), were women. Regarding education level, the majority, 52.8% (86), had a bachelor’s degree. About three-quarters of the respondents, 118 (72.4%), had a mean working experience of 7.49 years and a standard deviation (SD) of ± 3.101 years. In addition, less than half, 41.1% (67) of the study respondents attained training on sedation (Table 1).

The findings highlight that among 163 study respondents involved in this study, more than half, 68.1% (111), had adequate knowledge of sedation of critically ill patients. More than half of the respondents, 81.6% (133), were aware of any guidelines on sedation. Also, the majority of the study respondents, 92% (150), know the common indication for initiating sedation in ICU/HDU patients. The majority of study respondents, 94.5% (154), confirmed that the potential adverse effects of over-sedation in ICU patients are respiratory depression, hypotension, and prolonged mechanical ventilation. Furthermore, one hundred fifty-seven, 96.32% (157) know the Common risk associated with sedation in ICU/HDU among critically ill patients (Table 2).

After adjusting for all factors, gender remained significantly associated with knowledge of sedation. Nurses who were male were 38% less likely to have adequate knowledge of sedation compared to those who were female about sedation for critically ill patients (AOR = 0.377, 95% CI: 0.181-0.787, p = 0.009). Other factors, such as age, training, and educational level, were not significantly associated with knowledge of sedation in critically ill patients (Table 3)

The results highlighted that among 163 study respondents involved in this study, about three-quarters of the study respondents, 76.7% (125), had unfavourable attitudes about sedation of critically ill patients. The findings revealed that a small number of study respondents scored an average of 36.6% (1.83) on the item assessing how the nurse-patient ratio affects sedation management in the ICU during day and night shifts. Regarding having a feeling of being adequately trained to manage sedation protocols during both day and night shifts, they had a positive attitude with a score of 49.4 % (2.47). In addition, on items of Continuous pain assessment, which is necessary for effective sedation management in the ICU/HDU, the study respondents scored 47.0% (2.35) (Table 4)

After controlling for all factors, working position remained significantly associated with attitudes towards sedation of critically ill patients. The study found a significant association between training in sedation management and nurses’ attitudes. Nurses who had received training were two times more likely to demonstrate favourable attitudes compared to those without training (AOR = 2.553, 95% CI: 1.015–6.422, p = 0.046). Moreover, other factors like age, gender, education level, working experience, working institution, working ICU/HDU category, and training on sedation were not significantly associated with attitudes towards sedation of critically ill patients (Table 5).

The results revealed that among 163 study respondents involved in this study, only 33.1% (54) of study respondents used the standardized sedation scale (e.g., RASS, SAS) to assess sedation levels for critically ill patients. This highlighted that more than three-quarters, 81.6% (133) of study respondents had poor adherence practice of sedation to critically ill patients in ICUs/HDUS. Also, only 13.5% (22) of the study respondents use 1-2 hours to assess sedation levels during the shift (day/ night), whereas only 5.5% (9) of the nurses follow the sedation protocol guidelines. Furthermore, the Use of standardized sedation scale (e.g., RASS, SAS) to assess sedation levels were 33.1% (54) (Table 6).

After adjusting for all factors, age and year of experience in ICU/HDU remained significantly associated with the practices of sedation of critically ill patients. Nurses with an age range of 25 to 35 years were 34% less likely to exhibit poor adherence to sedation practice in critically ill patients compared to nurses whose ages were above 45 years (AOR = 0.344, 95% CI: 0.129-0.912, p = 0.032). Other factors like gender, working experience, education level, working position, and training on sedation were not statistically significantly associated with the practices of sedation of critically ill patients (Table 7).

## Discussion

The study evaluated nurses’ knowledge, attitudes, and practices regarding sedation in ICUs and HDUs in Dar es Salaam tertiary hospitals. Over two-thirds of nurses demonstrated adequate knowledge of sedation for critically ill patients. About three-quarters had an unfavourable attitude. More than three-quarters of all respondents demonstrated poor sedation adherence in this study. Females showed a significant predictor of sedation-related knowledge. Training in sedation management was associated with more positive attitudes toward sedation. Moreover, the age group between 25 and 35 years was significantly associated with sedation practices.

The adequate knowledge of sedation management observed in this study’s participants suggests a positive correlation with their reported frequent engagement in relevant training. This finding suggests that consistent, structured educational interventions have been effective in maintaining high standards among professionals responsible for patient sedation (24),(25). The findings further suggest that the majority of participants were relatively young, likely having recently completed their formal education. This demographic profile implies that their knowledge base is both current and actively maintained through recent academic exposure (12), (26). This demographic profile and training frequency collectively support the continuation of frequent, mandatory training programs to ensure sustained proficiency and reduce the incidence of management-related adverse events. This outcome is congruent with a study in Saudi Arabia and Pakistan, whereby 79.12 % and 67%of the respondents demonstrated adequate knowledge (27), (28). Conversely, a study in Ohio (USA) reported a significant lack of knowledge in sedation practice (25).

The unfavourable attitudes of our study participants towards sedation can be linked with high levels of burnout, emotional exhaustion, and moral distress. Often stemming from prolonged exposure to critically ill patients, ethical dilemmas, and resource constraints are known to impact nurses’ attitudes and engagement (2), (23). Furthermore, the key factors include a high workload and inadequate staffing, which lead to feelings of being overworked and result in missed nursing care(23). It can also be noted that the ICU is the environment where stressors such as increasing daily admissions, transfers, and deaths make participants feel overwhelmed, as reported in our study. This can be coupled with the low nurse-patient ratios, overworking, and poor life-work balance, leading to unfavourable attitudes among participants (17), (23).

Additionally, these findings corroborate a study reported in the U.S.A. that showed data indicating only 30.1%–43.2% of nurses acknowledge that peers’ attitudes influence their sedation practices (15). This mismatch with the findings of the study conducted in Canada data show 50% of Ottawa nurses reported positive attitudes toward sedation management.

Concerning the Sedation practices for critically ill patients in ICUs and HDUs, revealed that most respondents poorly adhered to sedation practices. The poor adherence to evidence-based sedation practices is largely due to a combination of institutional and professional barriers. It is evidenced that the absence of standardized, written protocols and a rigid hierarchy requiring physician orders inhibits the nurse autonomy necessary for implementing light sedation (29) & (7). Poor adherence to sedation practice can be described as the fact that Participants face a deep-seated fear of self-extubation or patient agitation perceived as unsafe. This is compounded by high workload and staffing shortages, leading to the use of easier-to-manage sedation methods. Thus, participants lack the authority and resources to consistently prioritize goal-directed and light sedation (30)& (27).

Furthermore, the absence of a protocol prevents, as reported in our study, integration of mandated assessments into charting systems, resulting in poor documentation and auditing failure, which ultimately makes it impossible for the unit to monitor adherence rates and implement effective quality improvements, as reported by (31). These findings are similar to the study conducted in Egypt, whereby 73.3% of participants showed poor adherence to sedative administration, highlighting protocol gaps in sedation practice (10). Contrary to a Norwegian study found 93% used RASS, reflecting strong adherence to standardized sedation assessment protocols (29).

The synthesis of these findings reveals a multifaceted challenge in sedation management. While knowledge levels are generally high among participants, well-trained nurses, attitudes, and practice adherence are undermined by systemic stressors and institutional limitations. Scientific evidence supports that regular, structured training programs, emotionally supportive work environments, and clear institutional protocols are essential to improving sedation outcomes. Addressing these factors holistically can enhance nurse autonomy, reduce adverse events, and promote consistent, evidence-based care in critical settings.

## Study limitations

Regardless of these strengths, the study is not without limitations. It was conducted within a specific geographic and institutional context, which may restrict the generalizability of its findings to other settings. The reliance on self-reported data introduces the potential for response bias, particularly when assessing subjective elements such as attitudes and practices. Furthermore, due to the cross-sectional design, the study is limited in its ability to infer causal relationships between variables.

## Conclusions

Despite having adequate knowledge of sedation practices, ICU nurses face persistent barriers in translating this knowledge into positive attitudes and consistent application. Barriers such as institutional culture, lack of support, or conflicting guidelines. Limited experience, high workloads, poor support, and inadequate training drive poor adherence and negative sedation attitudes. Education, leadership, and mentorship programs are crucial for closing sedation knowledge gaps and enhancing clinical practice. An encouraging and empowering clinical environment fosters improved sedation practices, enhancing patient safety and care quality in intensive care settings.

## Data Availability

All relevant data are within the manuscript and its Supporting Information files.

## Acknowledgements

We would like to thank the Muhimbili National Hospital in Dar es Salaam for the permission to conduct this study. We also extend our sincere gratitude to all the participants for their time, cooperation, and valuable insights, which greatly contributed to the development of this work.

